# Utilizing Experimental Cognitive Assessments and Machine Learning to Advance Prediction of Cognitive Impairment in Breast Cancer Survivors: A Preliminary Study

**DOI:** 10.1101/2025.11.14.25340009

**Authors:** Marc D. Rudolph, Keely A. Muscatell, Jessica R. Cohen

## Abstract

Up to 80% of women breast cancer survivors (BCS), particularly those treated with chemotherapy, report persistent cognitive impairment. Several meta-analyses and empirical studies assessing cancer and chemotherapy-related cognitive impairment (CRCI) in BCS have reported cognitive deficits across many general domains of cognition. Moreover, discrepancies often arise between objectively measured and self-reported impairment, potentially due, in part, to the use of non-specific neuropsychological assessments. Cognitive impairments faced by BCS may only be experienced by a portion of individuals due to the interaction of several contributing factors (e.g., age, fatigue, stress), although this possibility has not been assessed systematically. This study measured cognitive performance using multiple modalities: 1) self-report measures, 2) traditional neuropsychological assessments, and 3) experimental cognitive paradigms. Using machine learning, we generated models to distinguish BCS from healthy aging individuals without breast cancer, and to assess the relative predictive value of these three assessment modalities. We found that self-report measures of fatigue, distractibility, and stress were able to successfully differentiate BCS from healthy control participants. Further, a model combining these self-report measures with measures from experimental cognitive paradigms and neuropsychological assessments increased predictive accuracy; performance on cognitive paradigms ranked more important for prediction than performance on neuropsychological assessments. Although preliminary, results highlight the predictive utility of combining measures from several modalities and indicate that several distinct factors may contribute to CRCI. As the nature of CRCI is complex, this approach may help identify the best combination of assessments and performance metrics to measure subtle cognitive impairments in women BCS.

---

Breast cancer treatment is associated with persistent cognitive impairment in a substantial portion of breast cancer survivors (BCS) [1–6]. Though highly variable, both in terms of incidence and specific components of cognition impacted, up to 80% of breast cancer survivors report some form of cognitive impairment before, during, or after treatment with chemotherapy. Such impairment can negatively impact an individual’s quality of life and interfere with skills of daily living [7–9]. Given this impact, understanding the specific components of cognition affected, as well as the factors that predict those at risk for experiencing cognitive decline in BCS, is of paramount importance.

Several recent large-scale meta-analyses highlight that cognitive impairment among BCS is most common in the domains of memory, attention, and executive function, with additional reports of impaired processing speed [10–14]. However, findings from individual studies are inconsistent and there is a lack of consensus regarding both the presence and magnitude of cognitive impairment experienced by BCS. This may be in part because cognitive impairments experienced by BCS may be subtle and specific [15–19]. Indeed, inconsistencies in the literature may stem from the widespread use of neuropsychological assessments designed to detect broad, non-specific cognitive deficits in individuals with neurological disorders [20–22]. For example, traditional neuropsychological assessments used to measure executive function, such as the Delis-Kaplan Executive Function System (D-KEFS) Trail Making Test [23], simultaneously tap multiple executive and non-executive processes alike, including switching, inhibition, psychomotor speed, visual scanning, and sequencing [14]. Additionally, neuropsychological assessments condense complex cognitive processes into single summary scores. For example, performance on the Trail Making Test can be summarized as the number of errors committed or the total time required to complete the task. As such, it is not clear from overall Trail Making Test scores which specific cognitive processes are impaired. As with executive function, it is well-established that other high-level cognitive constructs such as attention and memory involve multiple components [24–27]. Thus, neuropsychological assessments are unable to reveal the specific cognitive processes that may be affected by cancer and its treatment. In contrast to neuropsychological assessments, experimental cognitive paradigms are designed to disentangle these subcomponents of cognitive processes within and across cognitive domains [6,22,28–31]. Additionally, such tasks include a large number of trials. Thus, as opposed to a single summary score, trial-wise metrics (i.e., accuracy, response time) that provide reliable measures of individual patterns of performance can be calculated [32–36]. The use of experimental cognitive paradigms may therefore improve detection of both general and specific subtle cognitive impairments experienced by some BCS [22,37,38].

Beyond simple detection of cognitive impairments faced by BCS, more research is also needed to understand why only a subset of individuals experience cognitive decline. It is currently unknown whether impairment is specific to certain types of cancer treatment (e.g., chemotherapy, endocrine therapy), or is a result of some combination of treatment and psychosocial, clinical, and demographic risk factors such as age, stress, sleep disturbances, or changes in mood [39–41]. Machine learning approaches that consider interactions between a set of factors in predicting an outcome, such as cognitive impairment, may improve our ability to understand such multifactorial relationships. Two seminal studies have used a combination of patient characteristics (e.g., age, cancer stage, depression, etc.) and metrics assessing brain network organization to identify whether this combination of factors was able to successfully predict cognitive impairment in BCS [42,43]. Machine learning tools were not only able to successfully distinguish individual BCS from healthy women [42] but were further able to predict future cognitive impairment at a one-year follow-up [43]. Cognition in these studies was either indirectly inferred via self-report or assessed with standardized summary scores from neuropsychological assessments. Thus, it remains unclear how these findings relate to impairment of specific cognitive processes.

In this preliminary study, we aimed to determine whether measures of memory, attention, and executive function from experimental cognitive paradigms could better differentiate BCS from healthy age-matched individuals without a history of breast cancer than neuropsychological assessments. Further, we assessed the role that demographics, psychosocial characteristics, and perceived cognitive failures may play in differentiating BCS from healthy participants beyond objective measures of cognitive functioning [44–46]. We built on prior machine learning work [2,22,47,48] by using a random forest ensemble method to identify the combination of features from experimental cognitive paradigms, neuropsychological assessments, and self-report measures that best distinguished BCS from healthy controls. We aimed to identify general and specific aspects of cognitive functioning, as well as demographic and self-reported health measures, that most contribute to accurate classification of BCS. We hypothesized that models including metrics from the experimental cognitive paradigms would be more accurate than models including neuropsychological assessments.

## METHODS & MATERIALS

### Participants

The current sample comprised 40 women ages 30-75: 20 women with primary, early-stage (I-IIIA) breast cancer who had completed chemotherapy within 1-12 months of their scheduled visit date (BCS group), and 20 female healthy control participants with no history of cancer or treatment with chemotherapy (see Table 1 for demographic characteristics). Groups were matched on age, ethnicity, and education. All participants were required to be proficient in English, have normal color vision, and have no history of neurologic or psychiatric disorders, with the exception of depression. Symptoms of depression are common contributing risk factors for CRCI [44–46] and were thus not considered exclusionary criteria and instead were assessed as predictors of interest. All BCS had completed adjuvant chemotherapy with or without concomitant endocrine therapy (tamoxifen or aromatase inhibitors). See *Supplementary Methods* and Table S1 for details on cancer diagnosis and treatment regiments.

**Table 1.** Group Demographic Characteristics.

| | BCS<br>(N=20) | Healthy Controls<br>(N=20) | $t/x^2$ | $p$ |
| --- | --- | --- | --- | --- |
| Age | 53.05 (9.89) | 49.65 (13.82) | 0.895 | .377 |
| Weight | 171.75 (52.82) | 160.00 (35.52) | 0.819 | .415 |
| WAIS Vocabulary | 47.11 (7.06) | 47.95 (9.90) | -0.302 | .765 |
| WAIS Matrix Reasoning | 18.53 (3.26) | 19.37 (7.34) | -0.457 | .652 |
| Race/Ethnicity |  |  | 3.949 | .557 |
| American Indian or Alaskan Native | 1 | 2 | ---- | ---- |
| Asian | 1 | 1 | ---- | ---- |
| Black/African-American | 3 | 1 | ---- | ---- |
| Native Hawaiian or Other Pacific Islander | 1 | 0 | ---- | ---- |
| White | 11 | 15 | ---- | ---- |
| Multiple Races | 3 | 1 | ---- | ---- |
| **Education** |  |  | 1.834 | .934 |
| Vocational or Training School | 1 | 1 | ---- | ---- |
| Some College (No Degree) | 1 | 1 | ---- | ---- |
| Associate Degree (2-year) | 1 | 1 | ---- | ---- |
| College Degree (4-year; BA or BS) | 5 | 6 | ---- | ---- |
| Some School Post-Degree (College or Professional) | 3 | 4 | ---- | ---- |
| Completed Master's Degree | 9 | 6 | ---- | ---- |
| Completed Doctoral Degree | 0 | 1 | ---- | ---- |

*Note:* Breakdown of demographic variables for all participants included in the current study. IQ was assessed using the Wechsler Adult Intelligence Scale (WAIS) Vocabulary and Matrix Reasoning subtests. Means, along with standard deviations in parentheses, are presented for continuous measures. Race/ethnicity and education levels are broken down separately for Breast Cancer Survivors (BCS) and healthy control participants. Group differences were assessed using t-tests for continuous measures and chi-squared tests for categorical measures. Abbreviations: BA = Bachelor of Arts; BCS = Breast Cancer Survivors; BS = Bachelor of Science; $t$ = Standard two-sample t-test; WAIS = Wechsler Adult Intelligence Scale; $\chi^2$ = chi-square test.

### Procedures & Measures

#### Overall Procedure

All participants completed a single session. The first portion of the session consisted of providing written informed consent and filling out demographic and health-related questionnaires on an iPad. Participants then completed a series of traditional neuropsychological assessments and experimental cognitive paradigms in counterbalanced order, with half of participants receiving neuropsychological assessments first and half receiving experimental cognitive paradigms first. Additionally, the administration order of individual tests within each assessment type was pseudo-randomized to control for fatigue, test difficulty, and trial order effects. Brief descriptions of all questionnaires and cognitive assessments are provided below.

See *Supplementary Methods* for further details regarding task design.

#### Questionnaires

Self-report questionnaires assessed a range of demographic and psychosocial factors that have been implicated in CRCI. In addition to demographics (e.g. age, weight, education, employment) and health history information not related to cancer or cancer treatment (e.g. non-cancer related medications), questionnaires assessed subjective cognitive functioning (Broadbent Cognitive Failure Questionnaire [CFQ; 49]), sleep quality (Pittsburgh Sleep Quality Index [PSQ; 52]), physical and mental fatigue (Multidimensional Fatigue Symptom Inventory Short Form [MFS; 51]), stress (Perceived Stress Scale [PSS; 53]), depressive symptoms (Beck Depression Inventory [BDI; 50]), and quality of life (36-Item Short-Form Health Survey [SF; 54]). The Functional Assessment of Cancer Therapy (FACT) [53] was administered to cancer survivors only to assess physical, functional, social, and emotional well-being post-treatment. Finally, a debriefing questionnaire (DQ) was administered to all participants to assess perceived difficulty with each of the three experimental cognitive paradigms (described below).

#### General Intelligence & Neuropsychological Assessments

General intelligence was estimated utilizing the Matrix Reasoning and Vocabulary subtests from the Wechsler Adult Intelligence Scale (WAIS) [23]. General aspects of memory, attention, and executive functioning were evaluated using assessments commonly employed in the CRCI literature including the Hopkins Verbal Learning Test-Revised (HVLT) [54], the D2 Test of Attention [55], and the Delis-Kaplan Executive Function System (D-KEFS) Trail Making and Color-Word Interference tests [23] respectively.

#### Experimental Cognitive Paradigms

To assess sub-processes within the general cognitive domains most affected by CRCI—memory, attention, and executive function—three experimental paradigms were selected: (1) the Relational and Item-Specific Encoding Task (RISE), (2) the Dot Pattern Expectancy Task (DPX), and (3) the Cued Cognitive Flexibility Task (CCF). These tasks target specific cognitive mechanisms and are designed to detect subtle impairments not captured by traditional assessments. Detailed descriptions of the RISE, DPX, and CCF tasks are provided below (see Figure 1), with additional information on task design, timing, and setup available in the *Supplementary Methods*.

**Figure 1.**
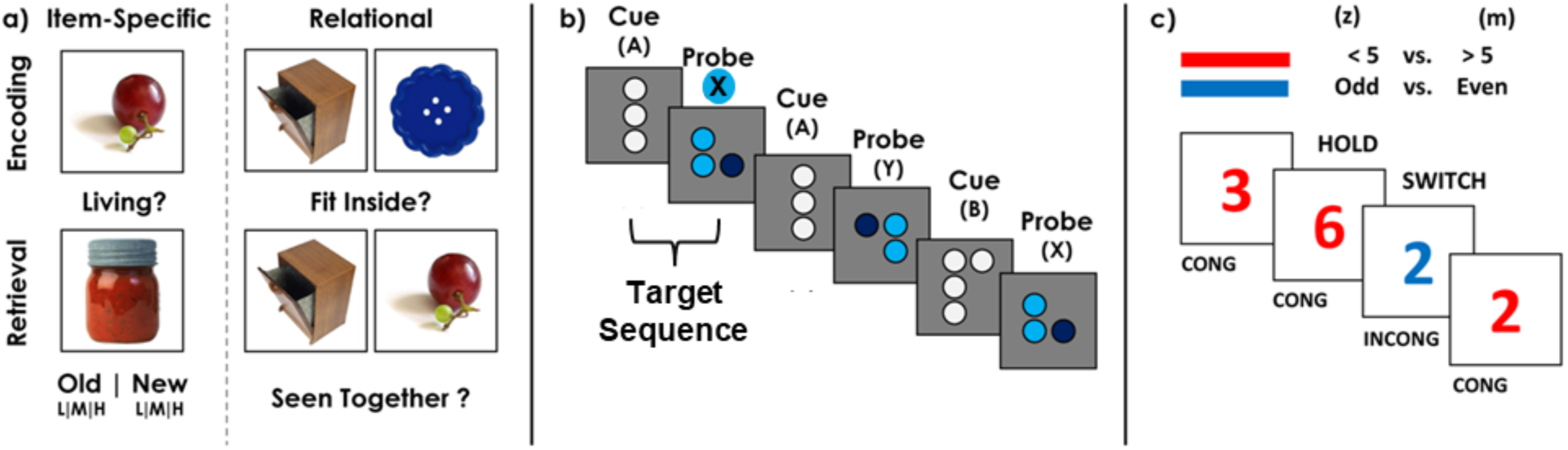
Experimental cognitive paradigms. a) The Relational and Item Specific Encoding (RISE) Task assessed memory encoding and retrieval for individual items (item-specific condition) and for associations between items (relational condition). During encoding, participants were asked to decide if an item was living or non-living (item-specific), or if one item could fit inside the other (relational). During retrieval, participants were asked to decide if an item was old or new (item-specific) or if a pair of items had been seen together previously (relational). In the item-specific retrieval condition, participants were additionally asked to rate how confident they were in their decision. b) The Dot Pattern Expectancy (DPX) Task assessed sustained and selective attention, working memory, response control, and processing speed. Participants viewed a series of dot patterns and were asked to identify whether a cue-probe sequence was a target sequence (pattern A followed by pattern X) or not. c) The Cued Cognitive Flexibility Task assessed cognitive flexibility, interference control, and implicit learning. Participants viewed a series of red or blue digits and were asked to decide if a number was less than or greater than five if red, or odd or even if blue. Trials could be hold (the color remained the same) or switch (the color changed). They could additionally be congruent (same response required for either the red or blue condition) or incongruent (different response required depending on color).

##### Relational and Item Specific Encoding (RISE) Task

The RISE is a two-part task [56] designed to distinguish between multiple stages of memory. It measures both memory encoding (part 1) and memory retrieval (part 2) for individual items (item-specific memory) and pairs of items (relational memory; Figure 1a). Part 1 consisted of three item-specific blocks with 12 trials each (36 trials total) and three relational blocks with 6 trials each (18 trials total), presented in alternating order. For item encoding, participants viewed a series of items one at a time and were asked to decide whether an item was living or non-living. For relational encoding, participants viewed a series of items in pairs and were asked if one item could fit inside the other. Part 2 consisted of one item retrieval block with 144 trials, and one relational retrieval block with 36 trials. For item retrieval, participants were asked to decide whether or not an item was seen previously during the item-specific encoding condition of part 1 and to rate their level of confidence in their decision (low, medium, high). For relational retrieval, participants were asked to decide whether or not a pair of objects had been seen together previously during the relational encoding condition of part 1. The RISE task used a set of 144 visual stimuli representing common everyday objects and scenes selected from a public repository of 2,500 color images (http://cvcl.mit.edu/MM/). By measuring accuracy during the different conditions, the RISE task distinguishes between episodic memory encoding and retrieval; by measuring response time variability during encoding, sustained attention is assessed; and by measuring mean response during encoding, processing speed is assessed.

##### Dot Pattern Expectancy (DPX) Task

The DPX task [57] was designed to distinguish between multiple cognitive components including sustained and selective attention, working memory, response control, and processing speed (Figure 1b). Stimuli consisted of a series of 3-5 white or blue dots arranged in one of four patterns (referred to here as A, B, X, and Y). Participants viewed a series of stimuli, one at a time, in cue-probe sequences. A and B were the two cue stimuli, while X and Y were the two probe stimuli. Participants were instructed to respond with a right (YES) button press to the target sequence (AX) and a left (NO) button press to all non-targets (AY, BX, BY). The target sequence occurred on 70% of trials, ensuring that was the prepotent response and increasing executive function and attention demands on the non-target sequences. The DPX task consisted of 4 blocks of 40 trials each (160 total). Sustained attention and processing speed are indexed be higher overall accuracy and faster response times. By contrasting accuracy and response times across the different trial types, this task has the ability to distinguish between selective attention (AY), working memory (AX|BX), and response control (AX|AY).

##### Cued Cognitive Flexibility (CCF) Task

The CCF task [58] was designed to distinguish between multiple components of executive function and learning (Figure 1c). In this task, participants viewed a series of randomly presented digits (0-9; excluding 5) and were cued by the digit’s color to make a judgment of either magnitude (red: greater or less than 5) or parity (blue: odd or even) with a left or right button press. Trials were either ‘hold’ (same judgment as the previous trial) or ‘switch’ (other judgment). Simultaneously, trials could be either ‘congruent’, whereby the stimulus presented required the same button press regardless of condition (i.e., 6 required a right button press for either magnitude or parity trials), or ‘incongruent’, whereby the stimulus presented required a different button across conditions (i.e., 7 required a right button press for magnitude and a left button press for parity). 50% of trials were congruent. This task was comprised of 3 blocks of 49 trials each (147 trials total). Each block had a different proportion of switch trials (25% [mostly hold], 50% [equal hold/switch], and 75% [mostly switch]) to determine whether implicit learning of switch frequency would influence task switching performance. Block order was counterbalanced across participants to ensure performance could not be attributed to individual differences in learning. By contrasting response times across trial types, this task distinguishes between two important sub-components of executive function, cognitive flexibility (performance decrement on ‘switch’ versus ‘hold’ trials) and interference control (performance decrement on ‘incongruent’ versus ‘congruent’ trials), and implicit learning (performance decrement in blocks with higher switch proportion).

#### Random Forest Classification

To assess how well BCS could be distinguished from healthy control participants based on cognitive function and other demographic, psychosocial, and clinical characteristics, the current study constructed supervised random forest classification models (Figure 2) using a set of 69 features from the questionnaires and cognitive assessments described above (see Table S2 in *Supplementary Methods* for a full list of features and summary statistics). Prior to random forest modeling, a series of data quality checks were performed to identify features exhibiting minimal variance, assess multicollinearity between features, and handle missing data to ensure classification models included informative and reliable predictors (see *Supplementary Methods* and *Supplementary Results*) resulting in a final set of 66 features.

**Figure 2.**
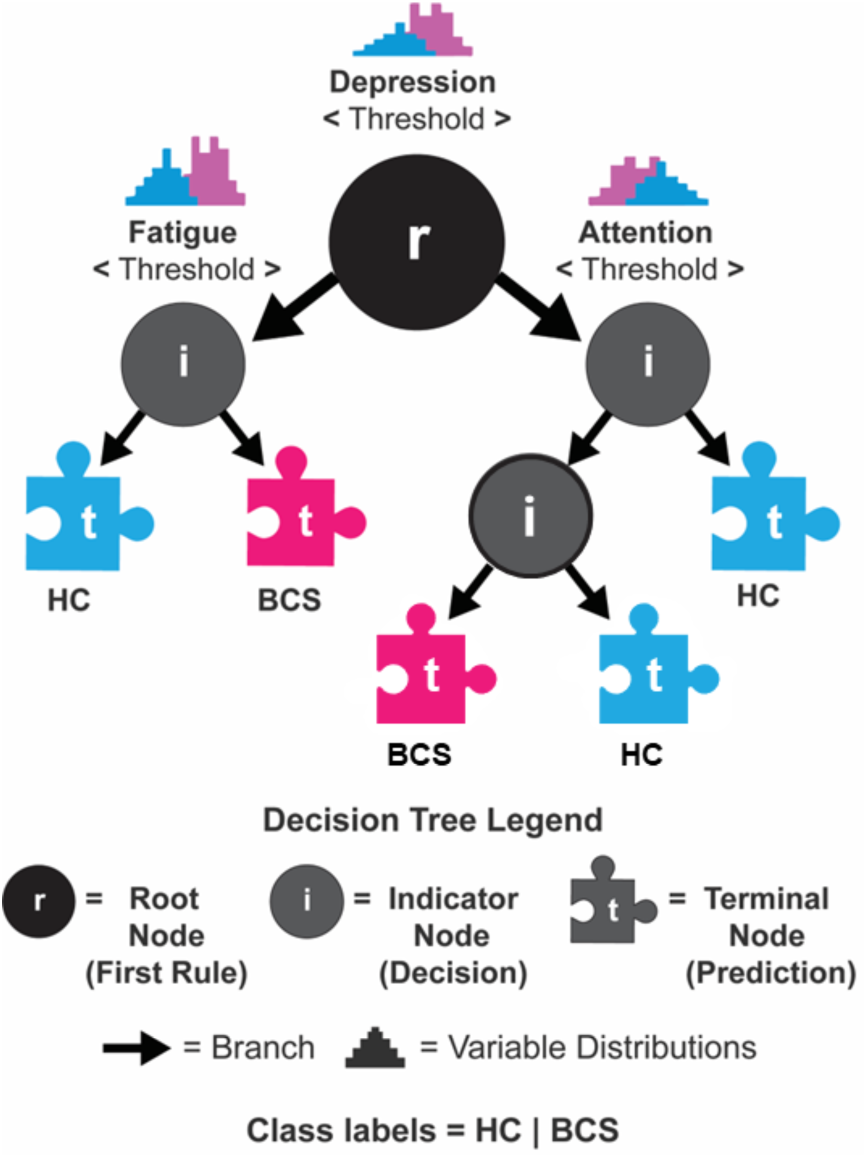
Simplified decision tree. This example depicts the basic components of a random forest decision tree. Starting from the top of a tree, the root node (r) represents the feature that best splits a set of observations into their respective classes (HC = healthy controls, BCS = breast cancer survivors). From the root node, a set of split observations is sent down their respective branch to the next decision point at an indicator node (i). This process is repeated until no more splits are possible and a decision is reached at a terminal node (t). At each terminal node, a final decision is made as to which class an observation belongs.

Next, five separate classification models were constructed. Our first three models utilized independent feature sets to establish whether experimental cognitive paradigms were better able to distinguish BCS from healthy control participants when compared to traditional neuropsychological assessments and self-report measures. Model 1 (EXP-COG) included only the 25 features from experimental cognitive paradigms, Model 2 included only the 20 features from neuropsychological assessments (NP), and Model 3 (SELF-REPORT) included only the 21 features assessing demographic, clinical, and psychosocial characteristics, along with perceived cognitive failures. Model 4 (FULL) included all 66 features.

A final, reduced model (Model 5; FINAL) included the top 20 ranked features across all assessments collected, including experimental cognitive paradigms, neuropsychological assessments, and self-report measures. The five random forest models were each constructed using 10-times repeated five-fold cross-validation [59–61]. On each fold of cross-validation, 2/3 of the data was randomly selected as a training set to generate classification models, which were then tested on the remaining 1/3. The Gini impurity index [62], which measures the probability that a randomly selected observation will be misclassified at a point in the decision tree, was used for model optimization and to inform feature selection. Permutation tests using randomly shuffled class labels (i.e., BCS vs. healthy control participant) were used to assess overall model significance. Finally, to isolate the most informative features, defined as those that contributed most to the accuracy of a given model, we computed variable importance based on the Mean Decrease in Impurity (e.g., Gini Importance; scaled from 0-100). We selected the top 20 features from the FULL model to include in the FINAL reduced model. From the EXP-COG, NP, and SELF-REPORT models, we selected the five most predictive features for visualization. We formally compared cross-validated models using several common performance metrics.

##### Model Comparisons

To formally compare the cross-validated models, we calculated several performance metrics. First, we calculated the area under the receiver operating curve (AUROC), a summary describing the probability of correctly identifying BCS cases (true positive; sensitivity) compared to those correctly classified as healthy control participants (true negative; specificity) across all iterations of cross-validation. As a complement to the AUROC, we calculated Informedness, which represents how accurate and balanced a set of predictions are for each class. Values range from -1 to 1 with zero representing chance performance and one indicating a perfect model. Additionally, we calculated the Brier score (0-1), which represents the difference between a set of predicted probabilities and perfect classification averaged across observations. A lower Brier score suggests greater precision (e.g., confidence) of a model’s prediction. Greater details of random forest modeling and model comparisons can be found in the *Supplementary Methods*.

### Post-Hoc Analyses (Supplementary Materials)

To determine whether time since completion of chemotherapy was related to the probability of being classified as BCS, we conducted a linear regression model testing for associations between time since treatment (dependent variable) and the probability of being classified as BCS using individual predictions from the FINAL random forest model. To permit comparisons with prior studies assessing CRCI, a series of post-hoc, two-sided independent-samples *t*-tests (alpha level set at .05) were conducted comparing BCS and healthy control participants on individual performance metrics (e.g. accuracy or mean RT from a given cognitive assessment). Descriptive statistics and uncorrected pairwise statistical comparisons are provided in Table S2 (*Supplementary Results*).

## RESULTS

### EXP-COG

A model including only metrics from the experimental cognitive paradigms had an accuracy of 50%, classifying 20 of 40 individuals correctly (9/20 BCS [45%]; 11/20 healthy controls [55%]) and was not significant compared to random chance (*p* = .509; Table 2). This model had a cross-validated AUROC of .59 (CI: .53-.65), a sensitivity of .53 (CI: .46-.60), and a specificity of .54 (CI: .47-.61). Informedness was .10 and the Brier-score was .245 (Table 2). The top 5 predictors (Figure S1a) included measures indexing RT on the RISE task (Relational and Item-Specific Encoding RT), sustained attention on the CCF task (Mostly Hold RT), and selective attention and working memory (AY & BX Target RT) on the DPX task.

**Table 2.** Cross-validated classification results.

| Performance Metric | EXP-COG | NP | SELF-REPORT | FULL | FINAL |
| --- | --- | --- | --- | --- | --- |
| ACC | .50 | .58 | .70 | .65 | .78 |
| AUROC | .59 | .69 | .81 | .75 | .87 |
| AUROC (CI) | .53-.65 | .64-.74 | .77-.85 | .70-.80 | .83-.91 |
| Sensitivity | .53 | .69 | .61 | .59 | .83 |
| Sensitivity (CI) | .46-.60 | .62-.75 | .54-.67 | .52-.65 | .77-.87 |
| Specificity | .54 | .61 | .78 | .76 | .82 |
| Specificity (CI) | .47-.61 | .54-.67 | .71-.83 | .70-.80 | .76-.86 |
| Informedness | .10 | .30 | .38 | .35 | .64 |
| Brier-score | .245 | .224 | .191 | .216 | .174 |

*Note:* Results for all classification models are presented together to promote efficient comparison. The results presented in the table indicate that the SELF-REPORT and FINAL reduced models produce the most accurate predictions overall. Abbreviations: ACC = Accuracy; AUROC = Area Under the Receiver Operating Curve; EXP-COG = Experimental Cognitive Paradigms; NP = Neuropsychological Assessments; CI = Confidence Interval (across all iterations of cross-validation).

### NP

A model including only metrics from the neuropsychological assessments had an accuracy of 58%, classifying 23 of 40 individuals correctly (12/20 BCS [60%]; 11/20 healthy controls [55%]) and was not significant compared to random chance (*p* = .091; Table 2). This model had a cross-validated AUROC of .69 (CI: .64-.74), a sensitivity of .69 (CI: .62-.75), and a specificity of .61 (CI: .54-.67). Informedness was .30 and the Brier-score was .224 (Table 2). The top 5 predictors (Figure S1b) included measures indexing concentration and errors on the D2 Test of Attention (Concentration Performance, Omission Errors), errors on the D-KEFS color-word inhibition test (Inhibition Errors, Switching Errors), and Immediate Recall on the HVLT.

### SELF-REPORT

A model including only metrics from self-report assessments had an accuracy of 70%, classifying 28 of 40 individuals correctly (13/20 BCS [65%]; 15/20 healthy controls [75%]) and was significant compared to random chance (*p* = .009; Table 2). This model had a cross-validated AUROC of .69 (CI: .64-.74), a sensitivity of .69 (CI: .62-.75), and a specificity of .61 (CI: .54-.67). Informedness was .30 and the Brier-score was .224 (Table 2). The top 5 predictors (Figure S1c) included measures indexing self-reported levels of fatigue (Total Fatigue on the SF), stress (Total Perceived Stress on the PSS), Distractibility on the CFQ, perceived difficulty on the CCF task (CCF Difficulty on the DQ), and age.

### FULL

The model including all available metrics had an accuracy of 65%, classifying 26 of 40 individuals correctly (12/20 BCS [60%]; 14/20 healthy controls [70%]) and was significant compared to random chance (*p* = .005; Table 2). This model had a cross-validated AUROC of .75 (CI: .70-.80), a sensitivity of .59 (CI: .52-.65), and a specificity of .76 (CI: .70-.80). Informedness was .35 and the Brier-score was .216 (Table 2). The top 5 predictors (Figure S1d) included measures indexing RT and accuracy on the RISE task (Relational Encoding RT, Item-Specific Encoding ACC), self-reported levels of fatigue (Total Fatigue on the SF), self-reported Distractibility on the CFQ, and working memory from the DPX task (BX Target RT).

### FINAL

A model comprised of the top 20 predictors selected from the FULL model had an accuracy of 78%, classifying 31 of 40 individuals correctly (15/20 BCS [75%]; 16/20 healthy controls [80%]) and was significant compared to random chance (*p* < .0001; Figure 3a; individual predictions are displayed in Figure 3b; Table 2). This model had a cross-validated AUROC of .87 (CI: .83-.91), a sensitivity of .83 (CI: .77-.87), and a specificity of .82 (CI: .76-.86). Informedness was .64 and the Brier-score was .174 (Table 2). All predictors in this model were preselected based on prior model performance and thus contribute to better classification of group status (BCS versus healthy control participants). The top 5 predictors (Figure 3c) included RT on the RISE task (Relational Encoding RT), self-reported levels of fatigue (Total Fatigue on the SF), self-reported Distractibility on the CFQ, selective attention on the DPX task (AY Target RT), and sustained attention on the CCF Task (Mostly Hold RT).

**Figure 3.**
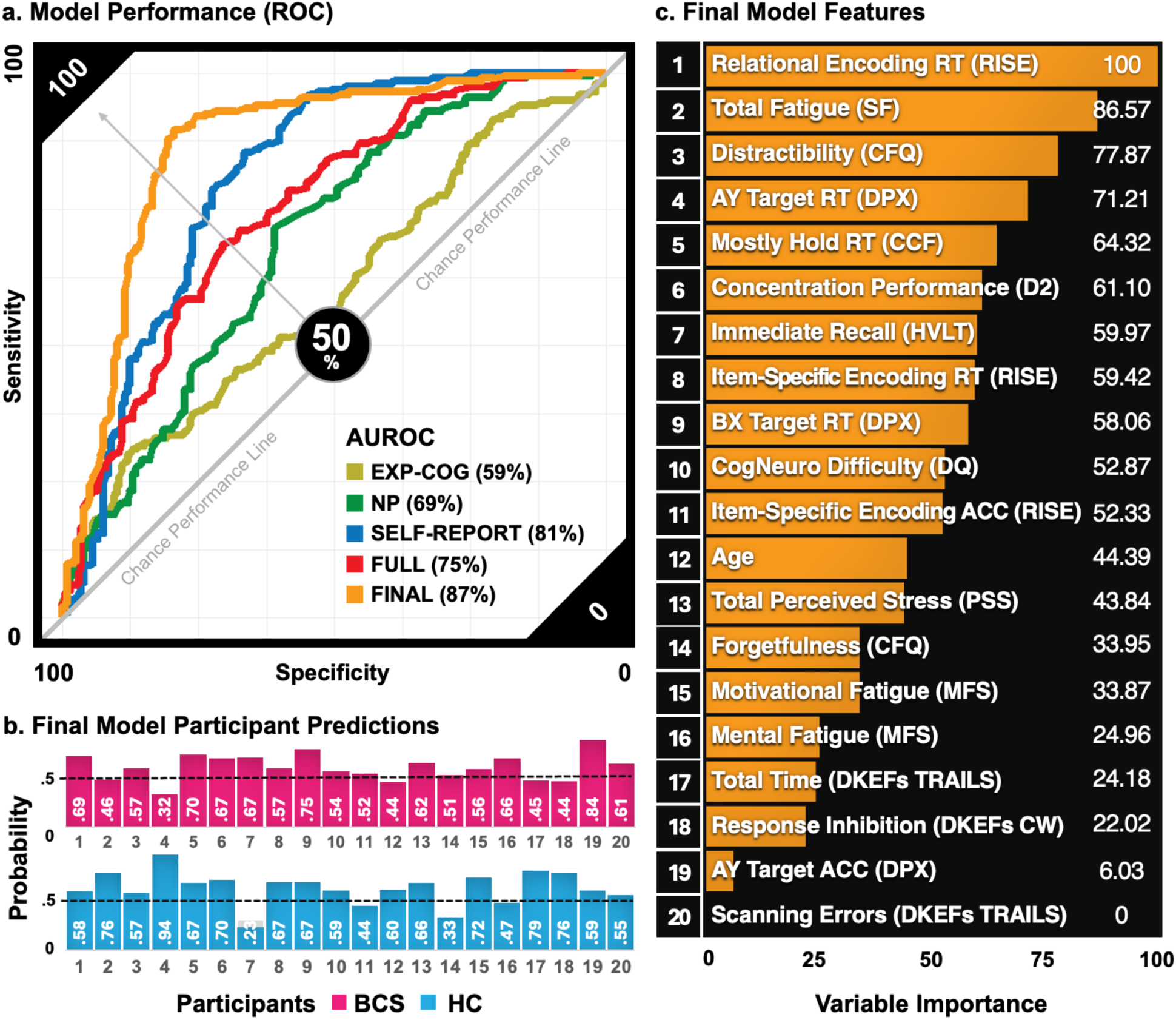
Model performance, variable importance scores, and individual predictions. a) ROC curves of all 5 models (EXP-COG, in gold; NP, in green; SELF-REPORT, in blue; FULL, in red; FINAL, in orange). AUROC demonstrates superior performance of the final model. b) Individual FINAL model predictions (e.g., the probability of being correctly classified as a BCS [top row, pink] or healthy control [bottom row, blue]). Predictions used a default binary decision threshold (e.g., 50%; random chance; dashed line). c) Variable importance scores show prediction was driven by a combination of metrics assessing general and specific aspects of attention, memory, and executive functioning, and self-report measures of fatigue, distractibility, and stress.

### Model Comparisons

As expected, the FINAL model, which was a reduced model that included the 20 most predictive variables from the FULL model, had the highest overall accuracy and overall provided the best sensitivity and specificity compared to all other models. The FINAL model was significantly more accurate (correcting for multiple comparisons) as compared to the EXP-COG, NP, and FULL models (EXP-COG: *t*(49) = 10.22, *p* < .001, *p-adj* < .001; NP: *t*(49) *= 7.34, p < .001,* , *p-adj* < .001; FULL: *t*(49) *= 5.85, p < .001, p-adj* < .001). Classification performance was not significantly different between the FINAL and SELF-REPORT models (*t*(49) = 2.31, *p* = .025, *p-adj* = .252). The SELF-REPORT model performed significantly better than the EXP-COG (*t*(49) = 6.53, *p* = < .001, *p-adj* = < .001) and NP (*t*(49) = 3.34, *p* = .002; *p-adj* = .016) models. Finally, the EXP-COG and NP models were not significantly different from one another (*t*(49) = -2.71, *p* = .010, *p-adj* = .091).

### Post-Hoc Analyses

First, we tested whether time since treatment was related to prediction accuracy on the FINAL model. We found no effect of time since treatment on individual class predictions (*R*^2^ = 0.086; *p* = .719). Next, we statistically compared performance on experimental cognitive paradigms, neuropsychological assessments, and self-report measures between BCS and healthy control participants. We observed that approximately 17% of features assessed (12 of 69) were significantly different between groups (*p* <= .05, *uncorrected*; see Table S2 in *Supplementary Results*). Three measures from the experimental cognitive paradigms were significantly different between BCS and healthy control participants. Specifically, BCS had slower response times on DPX BX trials indexing working memory and on the RISE memory encoding condition for both the item and relational conditions. Three measures from the neuropsychological assessments were significantly different between BCS and healthy control participants. Specifically, BCS had lower D2 concentration performance indexing attention, lower HVLT discrimination performance, and greater D-KEFS Trail-Making set-loss errors indexing cognitive flexibility. Finally, six self-report measures were significantly different between BCS and healthy control participants. Specifically, total symptoms of depression (BDS), total perceived stress (PSS), emotional and mental fatigue (MFS), and forgetfulness and distractibility (CFQ) were all greater in BCS as compared to healthy control participants.

## DISCUSSION

CRCI is multifaceted; multiple risk factors (e.g. age, genetics, stress, fatigue, physical and mental health) have been shown to alter the magnitude and trajectory of CRCI in various cognitive domains [1,63–65]. This preliminary study sought to capture the breadth of characteristics that could contribute to CRCI and determine the probability that BCS within 1-12 months of treatment with chemotherapy could be accurately distinguished from healthy control participants using a set of demographics, clinical, and psychosocial variables, as well as multiple assessments of cognitive functioning. Overall, results suggest that a combination of features from all modalities most accurately distinguished BCS from healthy control participants. Features that most contributed to accurate classification included: (1) measures from the experimental cognitive paradigms, particularly as related to memory (RISE) and attention (DPX); (2) self-reported measures of fatigue, distractibility, and stress; (3) measures from neuropsychological assessments indexing general aspects of attention (D2 Test of Attention) and response inhibition (D-KEFS CW); and (4) age.

When assessing models comprised of only one modality (experimental cognitive paradigms, neuropsychological assessments, or self-report assessments of psychosocial and cognitive functioning), only the model containing self-report measures successfully distinguished between BCS and healthy controls. Consistent with prior studies, elevated levels of fatigue and stress were among the top-ranked features that successfully predicted group membership [44,45,66–70]. This was also the case for models that additionally included measures indexing performance on experimental cognitive and neuropsychological assessments (i.e., the FULL and FINAL models). It is relatively well-established that psychosocial factors, such as mood and fatigue, tend to differ between patient and non-patient groups [39–41]. As expected, cross-validated performance of the SELF-REPORT model was comparable to the FULL model that additionally included cognitive assessments (cross-validated performance [AUROC]: SELF-REPORT=81%; FULL=75%). In contrast to the SELF-REPORT model, models composed of only cognitive assessments did not distinguish between BCS and healthy control participants relative to random chance. This was true both for the experimental cognitive paradigm (EXP-COG) and the neuropsychological assessment (NP) models. Contrary to expectations, these models performed significantly worse than all other models and relatively similarly to each other. This is similar to prior work that has found differences between BCS and healthy controls on measures assessing general aspects of attention, memory, and executive functioning, with several meta-analyses noting inconsistent findings across studies [11–13,71]. Thus, more work in larger samples is needed to identify the set of cognitive processes most impacted by cancer treatment and whether they have predictive utility over psychosocial measures or measures of self-reported cognitive difficulties.

Despite the lack of significant predictions of group membership in the EXP-COG and NP models, when identifying the top-performing features across all domains in the FULL and FINAL models, specific cognitive measures emerged as important for successful prediction. Notably, the most important feature in both the FULL and FINAL models was from the RISE experimental cognitive paradigm assessing memory performance. Altogether, two measures assessing encoding and retrieval performance on the RISE, two measures assessing selective attention and working memory on the DPX, and a single measure assessing sustained attention from the CCF were among the top 20 performing features in both the FULL and FINAL models. It is important to note that while measures from the experimental cognitive paradigms were ranked as more important than measures from the neuropsychological assessments, there were neuropsychological assessment measures that were ranked highly as well: most consistently, concentration performance on the D2 Test of Attention. While immediate recall performance on the HVLT and inhibition errors on the D-KEFS CW were also among the top ranked features in the FULL model, they were ranked 7^th^ and 18^th^ in the FINAL model, respectively. Together, this may indicate that even though the EXP-COG and NP models performed similarly, individual features from experimental cognitive paradigms are better able to distinguish BCS from healthy control participants when combined with self-report measures.

The RISE task appears to be particularly important for capturing differences between groups. However, studies conducted in both BCS [72] and in aging individuals without cancer [73], have found that difficulties in memory retention and recall may stem from impairments in attention. Here, consistent with elevated levels of self-reported distractibility and forgetfulness, and the importance of DPX measures assessing attention for group classification, deficits in sustained and selective attention observed among BCS may negatively impact memory encoding, which in turn would alter subsequent memory retrieval. In exploratory post-hoc analyses assessing group mean differences on specific measures, we observed that BCS had reduced concentration as assessed by the D2 Test of Attention (Table S2, *Supplementary Material*), supporting the idea that cognitive difficulties in BCS may emerge from impairments in attention. We additionally found that BCS had slower response times, on average, in the working memory condition of the DPX and in the item encoding condition of the RISE task, as well as more set-loss errors on the D-KEFS Trail-Making assessments. More work is needed to assess whether attention impairments are the root of objectively measured and subjectively reported CRCI.

### Limitations

Recruitment for this study was suspended March 1st, 2020, due to growing concerns over COVID-19 and the safety and well-being of our community. As a small pilot study, our findings must be taken with caution. First, although the random forest is one of the most stable machine learning algorithms [74], it is not immune to small sample issues. Specifically, the stability of model predictions and predicted class probabilities generated using small samples may not be reliable, regardless of cross-validation strategy, and can strongly depend on the characteristics of a given dataset [75]. We note, however, that previous studies assessing CRCI have successfully applied machine learning algorithms using similar sample sizes [43,76]. Second, specific characteristics of the current sample limit generalizability of our findings. The sample was comprised primarily of BCS who were HER2-(PR+/ER+/HER2-) or triple-negative (PR-ER-/HER2-; also see Table S1). HER2-positive tumors may be more aggressive and may confer additional risk for experiencing CRCI [77–79]. Additionally, all participants were fairly well-educated (85% had at least a 4-year degree) and the sample was mostly white/Non-Hispanic (65%, see Table 1). Lower levels of education have been associated with greater cognitive impairment in older non-white women BCS [80]. Thus, this sample may not include individuals who are at the greatest risk of experiencing CRCI. To address cancer-related health disparities and fully understand CRCI, it is critical that future studies pursue diverse samples. Third, participants practiced the experimental paradigms several times immediately prior to testing to ensure comprehension of task instructions, which may have resulted in artificially high baseline levels of performance, limiting our ability to detect intra- and interindividual variability across tasks. Finally, as a cross-sectional study it is not possible to assess the trajectory of cognitive impairment for a given individual, nor determine if and how baseline levels of cognitive ability before treatment impact the present results. However, consistent with prior work assessing short-term effects of CT on cognition [12,81–83], we found no association between time-since-treatment and the probability of being classified as BCS.

#### Conclusion

Previous research highlights that cognitive difficulties that negatively impact an individual’s quality of life and interfere with skills of daily living can be subtle yet have the potential to contribute to serious long-term impairment among BCS [15–19]. Understanding the precise nature of cognitive impairment in BCS is a critical step toward developing targeted early-intervention efforts aimed at minimizing the negative impacts of cognitive decline on the quality of life of breast cancer survivors [84]. Although preliminary, the present results may highlight the utility of combining experimental cognitive paradigms that permit an accurate decomposition of an individual’s performance across subcomponents of memory and attention with self-report measures that index physical and mental well-being. As the nature of CRCI is complex, this approach may help identify the best combination of assessments to objectively measure cognitive impairment in BCS. To further improve the accuracy, reliability, and specificity of models predicting which individuals are more at risk for experiencing short and long-term cognitive deficits, future longitudinal work can incorporate neurobiological markers (e.g., inflammatory cytokines, brain structure and function).

## Data Availability

All data produced in the present study are available upon reasonable request to the authors.

## Disclosure Statement

The authors have no competing interests to declare.

## Authorship Contribution Statement

**Marc D. Rudolph:** Conceptualization, Formal analysis, Methodology, Analytical Approach, Visualization, Writing - original draft, Writing - review & editing. **Keely A. Muscatell:** Conceptualization, Funding acquisition, Methodology, Resources, Supervision, Writing - original draft, Writing - review & editing. **Jessica R. Cohen:** Conceptualization, Funding acquisition, Methodology, Resources, Supervision, Writing - original draft, Writing - review & editing.

## Acknowledgments

This research was supported by a Tier 1 Pilot Award from the Lineberger Comprehensive Cancer Center at the University of Chapel Hill. We thank Dr. Elizabeth C. Dees for her consultation and help with recruitment and all UNC Breast Center providers within the Lineberger Comprehensive Cancer Center for referring patients. We also thank all participants for taking the time to complete our study.

## Disclosure Statement

The authors have no competing interests to declare.

## Notes

### Competing Interest Statement

The authors have declared no competing interest.

### Author Declarations

All procedures for this study were reviewed and approved by the Institutional Review Board at the University of North Carolina at Chapel Hill.

