## Supplemental Material for "Utilizing Experimental Cognitive Assessments and Machine Learning to Advance Prediction of Cognitive Impairment in Breast Cancer Survivors: A Preliminary Study"

### SUPPLEMENTARY METHODS

#### Tumor and Treatment-Specific Sample Characteristics

Participants with breast cancer were exposed to one of two commonly-used chemotherapy regimens: 1) 12 weeks: 4 cycles docetaxel and cyclophosphamide every 3 weeks; 2) 16 or 20 weeks: 4 cycles doxorubicin and cyclophosphamide every 2 weeks, followed by 4 cycles of paclitaxel either every 2 weeks or weekly for 12 cycles. The majority of BCS (N = 15) were progesterone (PR) and estrogen (ER) receptor positive and human epidermal growth factor receptor 2 (HER2) negative (PR+/ER+/HER2-). Three BCS were triple-negative (PR-ER-/HER2-) and two BCS were PR positive and ER and HER2 negative (PR-ER-/HER2-). Cancer stage and treatment plans were confirmed via electronic medical records in compliance with all Health Insurance Portability and Accountability Act of 1996 (HIPAA) regulations. See Table S1 for tumor and treatment-specific patient characteristics.

#### Experimental Cognitive Neuroscience Tasks

***Relational and Item Specific Encoding Task:*** The RISE is a two-part task [1] designed to distinguish between multiple stages of memory. It measures memory encoding and memory retrieval for both individual items (item memory) and pairs of items (relational memory). For a task overview and depiction, see *Main Text* (Experimental Cognitive Paradigms and Figure 1a). For item-specific encoding, participants viewed a series of items one at a time. Each item was presented for a duration of 2s with a 1s inter-trial interval between stimulus presentations. During the 2s stimulus presentation, participants were asked to decide whether or not an item was "living" or "non-living." Participants indicated their decision with a 2-button response (1=Yes; 2=No) using the index and middle fingers of their right hand. During the inter-trial interval, a white fixation cross was presented at the center of the screen. For relational encoding, participants viewed a series of items in pairs. Pairs of stimuli were presented for 4s each with a 1s inter-trial interval between presentation of the stimuli. During the 4s stimulus-pair presentation, participants were asked to decide whether one item could fit inside the other. Participants indicated their decision with a 2-button response (1=Yes; 2=No) using the index and middle fingers of their right hand.

Part 1 (encoding) consisted of 6 blocks: 3 item-specific and 3 relational. Blocks were presented in alternating order, beginning with an item-specific block. Before each block, instructions were presented for 3s to remind participants of the condition of the upcoming block. In the item-specific encoding blocks, each block contained 12 trials of one stimulus each (36 items in total). Relational encoding blocks contained 6 trials of two stimuli each (18 stimulus pairs in total). Stimulus pairs were presented adjacent to each other. For all conditions, participants were asked to respond as quickly and as accurately as possible. ***Training:*** Participants completed a practice run of 5 trials each for both the item-specific and relational encoding condition to ensure they understood the directions and knew which keys to press. Stimuli used during the practice were not repeated during the main task.

Part 2 (retrieval) was comprised of 2 blocks, 1 for the item-specific retrieval condition and 1 for the relational retrieval condition (*Main Text* Figure 1a). In the item-specific retrieval condition participants were asked to decide whether or not an item was seen previously during Part 1 and to simultaneously rate their level of confidence in their decision (low, medium, high). Specifically, participants marked an item as old (seen in Part 1) via a left-hand response and pressed one of three keys to indicate their level of confidence in their decision (a=low; s=medium; d=high). A right-hand response marked an item as new (not seen during the task) via a right-hand response using one of the three keys to indicate their level of confidence (j=high; k=medium; l=low). Stimuli were presented for a maximum of 10s, however, each trial ended 1.5s after a participant made a response. During the inter-trial interval, a white fixation cross was presented at the center of the screen for 1s. The item-specific retrieval block contained 144 trials (72 "new" visual items intermixed with 72 "old" visual items from Part 1, randomly intermixed). Before each block, instructions were read aloud to a participant. In the relational retrieval condition, participants were asked to decide whether or not a pair of objects had been seen together during the relational encoding condition in part 1. There was not a confidence judgment for this condition. Stimuli were presented for a maximum of 10s, during which participants responded with a 2-button response (1=Yes; 2=No) using the index and middle fingers of their right hand. Again, each trial ended 1.5s after a participant made a response and was followed by a fixation cross for 1s. The relational retrieval block contained 36 trials. Trials consisted of 18 randomly intermixed stimulus pairs from the relational encoding condition in part 1 and 18 new stimulus pairings (items previously seen in the relational encoding condition but not previously paired together). For all conditions, participants were asked to respond as quickly and as accurately as possible. ***Training:*** Participants completed a separate practice run before each retrieval condition. Each run consisted of 10 trials to ensure they understood the directions and knew which keys to press. Stimuli used during the practice were not repeated during the main task.

***Dot Pattern Expectancy Task (DPX)***. The DPX [2] is a modified AX continuous performance task that uses dot patterns instead of letters (see *Main Text* Figure 1b). The DPX was designed to distinguish between multiple cognitive components including sustained and selective attention, working memory, response control, and processing speed. In this task, participants viewed a series of stimuli that consisted of 3-5 white or blue dots arranged in a particular pattern. A total of four different patterns of dots were possible (referred to here as A, B, X, and Y). A and B were the cues, while X and Y were the probes. Each trial consisted of an initial cue, followed by a probe (AX, AY, BX, BY). Cue stimuli were presented for 1s, followed by a fixation cross for 2s. Probe stimuli were presented for .5s followed by a fixation cross for 1.9s. Participants were instructed to use the index and middle fingers of their right hand to respond with a YES ('k'key) button press to the probe of the target sequence (AX) and a NO ('j'key) button press to all cues and all probes from non-target sequences (AY, BX, BY). Thus, for the valid target (AX) sequence, participants were instructed to press the 'j' key to the valid cue 'A' and to press the 'k' key to the valid probe 'X'. For all other stimuli, participants were instructed to press the 'j' key. The target sequence occurred on 70% of trials, ensuring that was the prepotent response and increasing executive function and attention demands on the non-targets. AY and BX trials occurred on 12.5% of trials each. BY trials occurred on 5% of trials. On all trials, participants had 1.4s to respond to cue stimuli and .9s to respond to probe stimuli. Late responses were recorded but considered incorrect. After each allotted response window, 1 of 3 tones (buzz, knock, beep) was presented as audio feedback. These tones indicated whether or not a response was correct, incorrect, or late respectively. The DPX task consisted of 4 blocks of 40 trials for each of the 4 trial types (160 total). By contrasting accuracy and response times across the different trial types, this task has the ability to distinguish between sustained attention, selective attention, working memory, response control, and processing speed. *Training*: Before test administration, participants completed a series of training runs to ensure comprehension of task instructions. Training runs were identical to the test condition. Participants were told that their goal was to achieve an overall accuracy of 75% before moving on to the primary assessment. A maximum of 5 practice runs were administered. Participants were required to achieve an accuracy equal to or greater than 50% by the end of the 5 practice runs. This was the case for all participants.

***Cued Cognitive Flexibility Task (CCF).*** The CCF task [3] was designed to distinguish between multiple components of executive function and learning (see *Main Text* Figure 1c). In this task, participants viewed a series of randomly presented digits (0-9; excluding 5) and were cued by the digit's color to make a judgment of either magnitude (red: greater or less than 5) or parity (blue: odd or even) with a left or right button press using the index and middle fingers of their right hand. Trials were either 'hold' (same judgment as the previous trial) or 'switch' (other judgment). Simultaneously, trials could be either 'congruent', whereby the stimulus presented required the same button press regardless of condition (i.e., 6 required a right button press for either magnitude or parity trials), or 'incongruent', whereby the stimulus presented required a different button across conditions (i.e., 7 required a right button press for magnitude and a left button press for parity). 50% of trials were congruent. This task was comprised of 3 blocks of 49 trials each (147 trials total). Blocks differed in the proportion of switch trials presented (25%, 50%, or 75% switch). Block order was counterbalanced across participants to ensure task-switching performance could not be attributed to individual differences in stimulus-response learning. Stimuli were presented in random order for 1.2s. Following each stimulus, a fixation cross was displayed for a jittered inter-trial interval ranging from 1.25s to 2s (mean 1.6s). Inter-trial intervals were randomly pulled from an exponential distribution. By contrasting response times across trial types, this task distinguishes between two important sub-components of executive function: cognitive flexibility (performance decrement on 'switch' versus 'hold' trials) and interference control (performance decrement on 'incongruent' versus 'congruent' trials). ***Training:*** Prior to test administration, participants completed a series of training runs to ensure comprehension of task instructions. During training the proportion of switches was held constant at 50%. Participants were told that their goal was to achieve an overall accuracy level of 75% before moving on to the primary assessment. A maximum of 5 practice runs was administered. Participants were required to achieve an accuracy equal to or greater than 50% by the end of the 5 practice runs. This was the case for all participants.

**Random Forest Modeling**

Random forest models, cross-validation, and permutation tests were all carried out in R (R Core Team, 2020) using the *randomForest* [13], *caret* [14], and *rfUtilities* [18] packages, respectively. Random forest classification models were each constructed using 500 bootstrapped decision trees [4,5]. For each of the 500 bootstrapped decision trees, 70% of the available data was randomly selected as a training set to build the tree and the remaining 30% of data was used to test classification performance. Given our limited sample size, we additionally applied five-fold cross-validation, repeated 10 times (with replacement), to reduce overfitting and sampling bias and to ensure a sufficient number of observations were available for training (n=32) and testing (n=8) [6,7]. This has been shown to produce more reliable estimates of model performance as indexed using several common metrics described further below [6,8]. To handle missing data, we used the K-nearest neighbors algorithm to impute missing values by specifying *knnImpute* in the *preProcess* function within *caret*. Specifically, on each iteration of repeated cross-validation, knn imputation was performed exclusively on predictive features (x) within a given training set and applied to the test set [9–12]. The random forest algorithm randomly selects a subset of n features at each node in a decision tree (*mtry*). This parameter requires hyperparameter tuning to ensure optimal performance. For *mtry*, we used the grid search option in *caret* to select a subset of n possible predictors (minimum = 2; maximum = 8) at each split with a decision tree for a given model that resulted in the highest accuracy. The maximum number of predictors was set to equal the square root of the total number of possible features from the full model (n=66) [13,14]. Next, permutation tests were used to assess model significance in which the distribution of model misclassification rates, averaged across all independent test sets, was compared to a random null model. To generate empirical null models from the data, each step in the random forest process was repeated and class labels (i.e., group status: BCS vs. healthy control participants) were randomly shuffled. The number of permutations was set to 1000 [4,15–17].

***Model Comparisons.*** We compared the five random forest models on their respective ability to classify BCS from healthy control participants. Model sensitivity (true positive rate) and specificity (true negative rate) were calculated across a range of classification thresholds (0-1) using the receiver operating curve (ROC) [19]. The area under the ROC (AUROC) is a single numerical summary describing the probability of correctly identifying BCS cases (sensitivity) to those correctly classified as healthy control participants (specificity) for a given classification model. The distribution of AUROC, sensitivity, and specificity values, obtained by averaging performance across each iteration of cross-validation (10 repeats x 5 folds = 50 iterations), were formally compared between each model via a t-test using the *resamples* function in the *caret* R package [14], which by default applies a strict Bonferroni correction for multiple comparisons. Similar results were obtained when ROC curves were compared using the non-parametric Kolmogorov-Smirnov test and are thus not reported. As described in the main text, to complement the AUROC, we calculated both the Informedness and the Brier-score. Informedness is formally defined as a sum of the true positive rate (TPR; sensitivity) and the true negative rate (TNR; specificity) minus one (TPR+TNR-1). Positive values indicate higher true positive and true negative rates, indicating a lower number of false positives and false negatives respectively when predicting a target class (e.g., BCS). For example, a model with a sensitivity of 50% and specificity of 50% would thus yield an Informedness of 0 (.5 + .5-1) and would be unable to accurately distinguish between classes. Negative values indicate both an unbalanced and inaccurate model. Specifically, a model with a sensitivity of 50% and specificity of 0% would yield an Informedness of -.5 (.5 + 0-1) and would tend to classify observations into the wrong class at a rate greater than would be expected by chance. Finally, the Brier score considers the predicted probabilities used to classify observations into their respective classes. As noted in the main text, a lower Brier score suggests greater precision (e.g., confidence) of a model's prediction. Thus, a model that perfectly predicts each observation with 100% confidence would achieve a Brier score of 0. ROC metrics and plots were produced using the *caret* [14], *MLeval* [20], and *plotROC* [21] packages in R.

### Supplementary Results

***Quality control.*** As noted in the main text, prior to random forest modeling, a series of data quality checks were performed to assess multicollinearity between features, identify features exhibiting minimal variance, and handle missing data to ensure classification models included informative and reliable predictors. 69 possible features from experimental cognitive paradigms, neuropsychological assessments, and self-report measures (including age and weight) were examined. High multicollinearity may lead to bias during the random sampling process as similar features that are important for classification are likely to be overrepresented due to repeated selection. Features that lack variability cannot be used to differentiate items into their respective classes and only serve to increase computational time. Thus, features exhibiting a high degree of correlation ($r \geq .85$) and/or having near-zero variance were excised from further analysis [22]. Excluded features included scores on the Beck Depression Inventory (BDI Total; highly correlated with total perceived stress from the Perceived Stress Scale [PSS]), self-reported behavioral inhibition from the Cognitive Failures Questionnaire (CFQ; highly correlated with forgetfulness from the Multidimensional Fatigue Scale [MFS]), and total items processed from d2 Test of Attention (highly correlated with the total number of errors from the same test). No features exhibited near-zero variance; thus, no other features were excluded from further analysis. Items not excluded during quality control analyses were used as predictive features to distinguish BCS from HC using random forest classification modeling. Finally, the dataset was inspected for missing values. A small portion (1.1%) of data points were missing at random. Missing data was observed for 4 of 40 participants (2 HC, 2 BCS) missing 13 (19%), 4 (6%), 2 (3%), and 9 data points respectively. Missing values were imputed during model training as described above.

**Post-hoc Analyses**

***Group Comparisons.*** A series of post-hoc, uncorrected, Welch two-sided independent t-tests (alpha level set at .05) were conducted comparing BCS and healthy control participants on individual performance metrics from experimental cognitive paradigms, neuropsychological assessments, and self-report questionnaires (see *Main Text:* Methods, Post-hoc Analyses). 12 of 69 variables (17%) exhibited group mean differences (3 from experimental cognitive paradigms, 3 from neuropsychological assessments, and 6 from self-report questionnaires; see Table S2). From the experimental cognitive paradigms, compared to healthy controls, BCS had slower response times in the working memory condition of the DPX (DPX BX Target RT; $t(36.64) = 2.04$, $p = .048$) and on the RISE task for both the item-specific encoding condition (RISE Item-Specific Encoding RT; $t(36.99) = 2.23$, $p = .032$) and the relational encoding condition (RISE Relational Encoding RT; $t(36.99) = 2.53$, $p = .016$). On neuropsychological assessments, BCS exhibited poorer attention and mental flexibility compared to healthy controls as indexed by lower concentration performance on the D2 Test of Attention ($t(32.35) = -2.05$, $p = .048$), lower discrimination performance on the Hopkins Verbal Learning Test (HVLT; $t(23.03) = -2.26$, $p = .034$), and a greater number of set-loss errors on the switching condition of the Delis-Kaplan Executive Function System (D-KEFS) Trail-Making test $t(19) = -2.67$, $p = .015$). From self-report questionnaires, total self-reported symptoms of depression measured by the Beck Depression Inventory (BDI), psychosocial stress measured by the Perceived Stress Scale (PSS), and both emotional and mental fatigue measured by the Multi-dimensional Fatigue Scale (MFS) were significantly higher in BCS than in healthy control participants (BDI Total: $t(31.64) = 2.98$, *p* = .005; PSS Total: *t*(29.29) = 2.27, *p* = .031; MFS Emotional: *t*(31.35) = 2.15, *p* = .040; MFS Mental: *t*(30.90) = 2.44, *p* = .021). Forgetfulness and distractibility as measured by the Cognitive Failures Questionnaire (CFQ) were also higher in BCS compared to healthy controls (CFQ Forgetfulness: *t*(36.38) = 2.43, *p* = .020; CFQ Distractibility: *t*(36.52) = 2.67, *p* = .011).

### Supplementary Discussion

With the exception of set-loss errors on the D-KEFS Trail-Making Test and discrimination on the HVLT, features indicating potential group mean differences were also important features for classifying BCS from healthy controls in both the FULL and FINAL models (see Results and Figures 3d and 4c in *Main Text*). Other features implicated in CRCI that did not have group mean differences (fatigue, sleep, age, etc.) were important for classifying individuals exhibiting some level of cognitive difficulty. These variables may help identify tasks and conditions for which there is a greater deal of individual cognitive variability within and between groups. Again, however, caution of overinterpreting is warranted given the lack of statistical power of this sample to assess group mean differences under the null-hypothesis testing framework. Findings may further highlight the importance of considering high-level interactions between both distinct and overlapping cognitive processes in heterogenous groups.

### Supplementary Figures & Tables

#### Table S1. Tumor and Treatment-Specific Sample Characteristics

| | | | | | |
|---|---|---|---|---|---|
| **a.** | **Time Since Treatment** | **1-3 mo** | **4-6 mo** | **7-9 mo** | **10-12 mo** |
| | | N = 8 | N = 5 | N = 4 | N = 3 |
| **b.** | **Treatment Regimen** | **ddAC** | **ddAC+PAC** | **PAC** | -- |
| | | N = 9 | N = 10 | N = 1 | -- |
| **c.** | **Receptor Type** | **(PR+/ER+/HER2-)** | **(PR+/ER-/HER2-)** | **(PR-/ER-/HER2-)** | -- |
| | | N = 15 | N = 2 | N = 3 | -- |
| **d.** | **Tumor Grade** | **Grade 1** | **Grade 2** | **Grade 3** | -- |
| | | N = 4 | N = 12 | N = 4 | -- |

*Note:* Breakdown of treatment information for breast cancer survivors. (a) All patients completed the study within 12 months of completion of treatment with CT. The majority of BCS: (b) received Dose-Dense Adjuvant Doxorubicin (ddAC), (c) were progesterone (PR) and estrogen (ER) receptor positive and human epidermal growth factor receptor 2 (HER2) negative (PR+/ER+/HER2-), and (d) had a tumor grade of 2. Abbreviations: + = Positive; - = Negative; ddAC = Dose-Dense Adjuvant Doxorubicin and Cyclophosphamide; ddAC+PAC = Dose-Dense Adjuvant Doxorubicin and Cyclophosphamide Plus Paclitaxel; ER = estrogen; HER2 = human epidermal growth factor receptor 2; PAC = Paclitaxel; PR = Progesterone

**Table S2. Summary statistics and group comparisons.**

| | Mean | | SD | | Median | | Skew | | Inference | |
|---|---|---|---|---|---|---|---|---|---|---|
| | **HC** | **BCS** | **HC** | **BCS** | **HC** | **BCS** | **HC** | **BCS** | ***t*** | ***p*** |
| EXPERIMENTAL COGNITIVE PARADIGMS | | | | | | | | | | |
| RISE Item-Specific Encoding ACC | 0.87 | 0.83 | 0.06 | 0.06 | 0.86 | 0.82 | -0.47 | 0.76 | 1.77 | .085 |
| **RISE Item-Specific Encoding RT** | **0.97** | **1.09** | **0.16** | **0.17** | **0.95** | **1.08** | **0.11** | **-0.16** | **-2.23** | **.032** |
| RISE Relational Encoding ACC | 0.76 | 0.77 | 0.10 | 0.11 | 0.72 | 0.78 | 0.06 | 0.30 | -0.12 | .907 |
| **RISE Relational Encoding RT** | **1.73** | **1.99** | **0.32** | **0.34** | **1.77** | **2.01** | **0.46** | **-0.91** | **-2.53** | **.016** |
| RISE Item-Specific Retrieval ACC | 0.92 | 0.92 | 0.04 | 0.04 | 0.92 | 0.92 | -0.87 | -0.47 | 0.26 | .799 |
| RISE Item-Specific Retrieval RT | 1.45 | 1.53 | 0.27 | 0.40 | 1.46 | 1.54 | 0.33 | 0.34 | -0.71 | .485 |
| RISE Relational Retrieval ACC | 0.84 | 0.80 | 0.12 | 0.12 | 0.85 | 0.78 | -0.19 | -0.08 | 0.95 | .348 |
| RISE Relational Retrieval RT | 2.09 | 2.43 | 0.45 | 0.66 | 1.99 | 2.21 | 0.83 | 0.57 | -1.93 | .062 |
| DPX Target AX ACC | 0.94 | 0.95 | 0.05 | 0.03 | 0.96 | 0.95 | -1.26 | -0.58 | -0.46 | .649 |
| DPX Target AX RT | 0.42 | 0.43 | 0.05 | 0.05 | 0.42 | 0.42 | 0.29 | 0.02 | -0.69 | .493 |
| DPX Target AY ACC | 0.72 | 0.69 | 0.26 | 0.22 | 0.78 | 0.73 | -0.75 | -1.61 | 0.34 | .733 |
| DPX Target AY RT | 0.52 | 0.57 | 0.09 | 0.06 | 0.53 | 0.57 | -0.59 | -0.74 | -1.98 | .056 |
| DPX Target BX ACC | 0.90 | 0.87 | 0.11 | 0.13 | 0.95 | 0.90 | -0.77 | -1.13 | 0.77 | .449 |
| **DPX Target BX RT** | **0.39** | **0.44** | **0.06** | **0.08** | **0.37** | **0.44** | **0.41** | **-0.29** | **-2.04** | **.048** |
| DPX Target BY ACC | 0.90 | 0.95 | 0.14 | 0.09 | 1.00 | 1.00 | -1.24 | -2.07 | -1.35 | .186 |
| DPX Target BY RT | 0.42 | 0.44 | 0.09 | 0.09 | 0.44 | 0.47 | -0.07 | -0.20 | -0.71 | .485 |
| CCF EQ ACC | 84.99 | 88.71 | 12.44 | 7.96 | 88.54 | 91.67 | -0.88 | -0.46 | -1.12 | .273 |
| CCF EQ Cost | 116.40 | 150.10 | 121.14 | 129.92 | 117.28 | 131.48 | 0.68 | 0.66 | -0.84 | .408 |
| CCF EQ RT | 917.37 | 916.08 | 104.57 | 144.74 | 907.87 | 891.97 | 0.61 | 1.85 | 0.03 | .975 |
| CCF MH ACC | 87.95 | 92.43 | 8.62 | 4.87 | 90.63 | 91.67 | -1.04 | 0.00 | -2.01 | .053 |
| CCF MH Cost | 154.72 | 202.66 | 148.10 | 115.44 | 142.27 | 181.91 | 0.43 | 1.14 | -1.13 | .266 |
| CCF MH RT | 870.54 | 848.32 | 101.25 | 84.31 | 874.82 | 852.29 | 0.31 | 1.46 | 0.75 | .460 |

| | | | | | | | | | | |
|---|---|---|---|---|---|---|---|---|---|---|
| CCF MS ACC | 85.96 | 90.35 | 13.62 | 6.45 | 87.50 | 91.67 | -1.50 | -0.87 | -1.30 | .205 |
| CCF MS Cost | 96.40 | 82.20 | 91.07 | 104.81 | 102.24 | 63.06 | -0.01 | 0.25 | 0.45 | .655 |
| CCF MS RT | 926.60 | 948.39 | 127.84 | 163.03 | 917.42 | 943.81 | 1.37 | 2.00 | -0.46 | .646 |
| NEUROPSYCHOLOGICAL ASSESSMENTS | | | | | | | | | | |
| WAIS Vocabulary | 47.95 | 47.11 | 9.90 | 7.06 | 50.00 | 48.00 | -2.25 | -0.17 | 0.30 | .765 |
| WAIS Matrix Reasoning | 19.37 | 18.53 | 7.34 | 3.26 | 20.00 | 20.00 | 1.44 | -0.69 | 0.46 | .652 |
| D2 Omission Errors | 26.10 | 28.05 | 21.55 | 25.11 | 23.00 | 17.50 | 1.25 | 0.63 | -0.26 | .794 |
| D2 Total Errors | 468.95 | 449.65 | 85.44 | 68.21 | 470.00 | 456.50 | 0.29 | -0.37 | 0.79 | .435 |
| D2 Total Items Processed | 495.00 | 477.55 | 91.75 | 77.10 | 480.50 | 477.50 | 0.33 | -0.38 | 0.65 | .519 |
| **D2 Concentration Performance** | **200.70** | **177.80** | **42.15** | **27.00** | **197.50** | **178.00** | **0.20** | **-0.23** | **2.05** | **.049** |
| HVLT Immediate Recall | 30.70 | 28.65 | 3.20 | 4.28 | 31.50 | 28.00 | -0.26 | 0.28 | 1.72 | .095 |
| HVLT Delayed Recall | 10.85 | 10.15 | 1.46 | 1.60 | 11.00 | 10.00 | -1.68 | -0.23 | 1.45 | .157 |
| HVLT Memory Retention | 97.15 | 96.00 | 8.82 | 10.47 | 100.00 | 100.00 | -1.77 | 0.17 | 0.38 | .709 |
| **HVLT Discrimination** | **11.90** | **11.40** | **0.31** | **0.94** | **12.00** | **12.00** | **-2.47** | **-1.17** | **2.26** | **.034** |
| D-KEFS CW Total Time | 395.21 | 380.35 | 110.65 | 81.13 | 368.00 | 360.00 | 1.82 | 0.88 | 0.48 | .637 |
| D-KEFS CW Response Inhibition | 50.95 | 49.75 | 15.35 | 6.89 | 45.00 | 49.50 | 0.49 | 0.59 | 0.31 | .758 |
| D-KEFS CW Response Inhibition Errors | 0.42 | 0.65 | 0.69 | 0.75 | 0.00 | 0.50 | 1.22 | 0.60 | -0.99 | .326 |
| D-KEFS CW Switching | 56.16 | 58.00 | 12.27 | 9.53 | 56.00 | 55.00 | 0.05 | 1.16 | -0.52 | .605 |
| D-KEFS CW Switching Errors | 0.89 | 1.05 | 1.20 | 1.32 | 1.00 | 1.00 | 2.03 | 1.36 | -0.39 | .702 |
| D-KEFS Trails Total Time | 514.37 | 462.90 | 233.58 | 96.52 | 464.00 | 462.50 | 3.32 | 2.52 | 0.89 | .382 |
| D-KEFS Trails Scanning Total Time | 0.16 | 0.20 | 0.37 | 0.52 | 0.00 | 0.00 | 1.73 | 2.35 | -0.77 | .447 |
| D-KEFS Trails Scanning Total Errors | 21.68 | 23.55 | 7.95 | 7.16 | 19.00 | 22.50 | 1.12 | 1.03 | -0.29 | .774 |
| D-KEFS Trails Switching Total Time | 66.32 | 70.05 | 18.00 | 27.08 | 64.00 | 61.50 | 0.47 | 1.52 | -0.51 | .614 |

| | | | | | | | | | |
|---|---|---|---|---|---|---|---|---|---|
| D-KEFS Trails Switching Sequence Errors | 0.32 | 0.15 | 0.58 | 0.49 | 0.00 | 0.00 | 1.50 | 2.94 | 0.96 | .344 |
| **D-KEFS Trails Switching Set-Loss Errors** | **0.00** | **0.35** | **0.00** | **0.59** | **0.00** | **0.00** | **0.00** | **1.30** | **-2.67** | **.015** |
| SELF-REPORT MEASURES | | | | | | | | | | |
| Age | 49.65 | 53.05 | 13.82 | 9.89 | 53.5 | 50 | 0.20 | -0.05 | -.895 | .377 |
| Weight | 160.00 | 171.75 | 35.52 | 52.82 | 146.5 | 163 | 0.50 | 0.61 | -.819 | .415 |
| **BDI Total** | **4.45** | **11.30** | **5.39** | **8.74** | **3.00** | **10.00** | **1.64** | **0.68** | **-2.98** | **.005** |
| **PSS Total Perceived Stress** | **11.20** | **16.75** | **5.22** | **9.62** | **11.00** | **16.50** | **0.14** | **0.24** | **-2.27** | **.031** |
| PSQ Sleep Quality | 1.95 | 2.15 | 0.94 | 1.04 | 2.00 | 3.00 | -0.26 | -0.55 | -0.64 | .528 |
| PSQ Sleep Duration | 7.95 | 10.70 | 5.13 | 5.01 | 7.00 | 11.50 | 0.54 | 0.03 | -1.71 | .095 |
| MFS Physical Fatigue | 2.05 | 4.50 | 2.93 | 4.82 | 1.00 | 2.00 | 2.01 | 1.07 | -1.94 | .061 |
| **MFS Emotional Fatigue** | **3.20** | **6.55** | **3.62** | **5.96** | **2.50** | **5.00** | **1.49** | **0.45** | **-2.15** | **.040** |
| **MFS Mental Fatigue** | **3.50** | **7.40** | **3.65** | **6.15** | **2.00** | **6.00** | **0.92** | **0.39** | **-2.44** | **.021** |
| MFS Motivational Fatigue | 14.50 | 12.15 | 4.01 | 7.05 | 15.00 | 14.00 | -0.76 | -0.11 | 1.30 | .205 |
| **CFQ Forgetfulness** | **11.00** | **14.70** | **4.28** | **5.30** | **10.50** | **14.50** | **0.56** | **-0.13** | **-2.43** | **.020** |
| **CFQ Distractibility** | **9.10** | **13.35** | **4.51** | **5.53** | **8.00** | **13.50** | **0.61** | **0.10** | **-2.67** | **.011** |
| CFQ Inhibition | 5.80 | 7.70 | 3.29 | 3.77 | 5.50 | 7.00 | 0.45 | 0.37 | -1.70 | .098 |
| SF General Scale | 72.50 | 71.31 | 20.29 | 19.96 | 75.00 | 75.00 | -0.46 | -0.46 | 0.19 | .853 |
| SF Physical Scale | 85.91 | 73.18 | 21.67 | 27.27 | 95.45 | 86.36 | -1.68 | -0.99 | 1.63 | .111 |
| SF Fatigue Scale | 57.50 | 56.50 | 22.80 | 29.20 | 67.50 | 55.00 | -0.83 | -0.11 | 0.12 | .905 |
| SF Emotional Scale | 80.80 | 73.00 | 12.56 | 22.73 | 84.00 | 84.00 | -1.44 | -0.77 | 1.34 | .189 |
| SF Social Scale | 84.38 | 78.75 | 24.29 | 24.37 | 100.00 | 81.25 | -1.46 | -0.73 | 0.73 | .469 |
| SF Pain Scale | 78.13 | 71.75 | 18.88 | 22.42 | 85.00 | 67.50 | -0.42 | -0.50 | 0.97 | .337 |
| DQ DPX Difficulty | 9.70 | 11.30 | 4.85 | 6.40 | 9.00 | 11.50 | 0.23 | 0.20 | -0.89 | .379 |
| DQ RISE Difficulty | 7.55 | 9.90 | 4.75 | 6.72 | 7.00 | 9.50 | 0.73 | 0.07 | -1.28 | .210 |
| DQ CCF Difficulty | 16.90 | 15.15 | 4.89 | 7.20 | 16.50 | 15.00 | 0.18 | -0.08 | 0.90 | .375 |

| | | | | | | | | | | |
|---|---|---|---|---|---|---|---|---|---|---|
| DQ Overall Difficulty | 11.15 | 12.10 | 4.39 | 6.18 | 13.00 | 12.50 | -0.97 | -0.05 | -0.56 | .579 |

*Note:* Descriptive statistics are provided for all variables considered for analysis in the current study from experimental cognitive paradigms, neuropsychological assessments, and self-report questionnaires. In addition, performance metrics from experimental cognitive paradigms and neuropsychological assessments and average scores from self-report questionnaires were statistically compared between healthy control participants and BCS using Welch two-sample independent t-tests. Values displayed in bold typeface reflect statistically significant group differences assessed via a standard Welch two-sample t-test ($p \le .05$; uncorrected).
*Abbreviations.* ACC = Accuracy; BDI = Beck Depression Inventory; CCF: Cueing Cognitive Flexibility Task; CFQ = Cognitive Failures Questionnaire; D-KEFS = Delis-Kaplan Executive Function System; DPX = Dot Pattern Expectancy Task; DQ = Debriefing Questionnaire; PSS = Perceived Stress Scale; PSQ = Sleep Quality Inventory; MFS = Multidimensional Fatigue Scale; RISE = Relational and Item-Specific Encoding Task; RT = Mean Response Time; SF = Short-Fatigue Scale; WAIS = Weschler Adult Intelligence Scale

**Figure S1. Cross-validated feature performance**

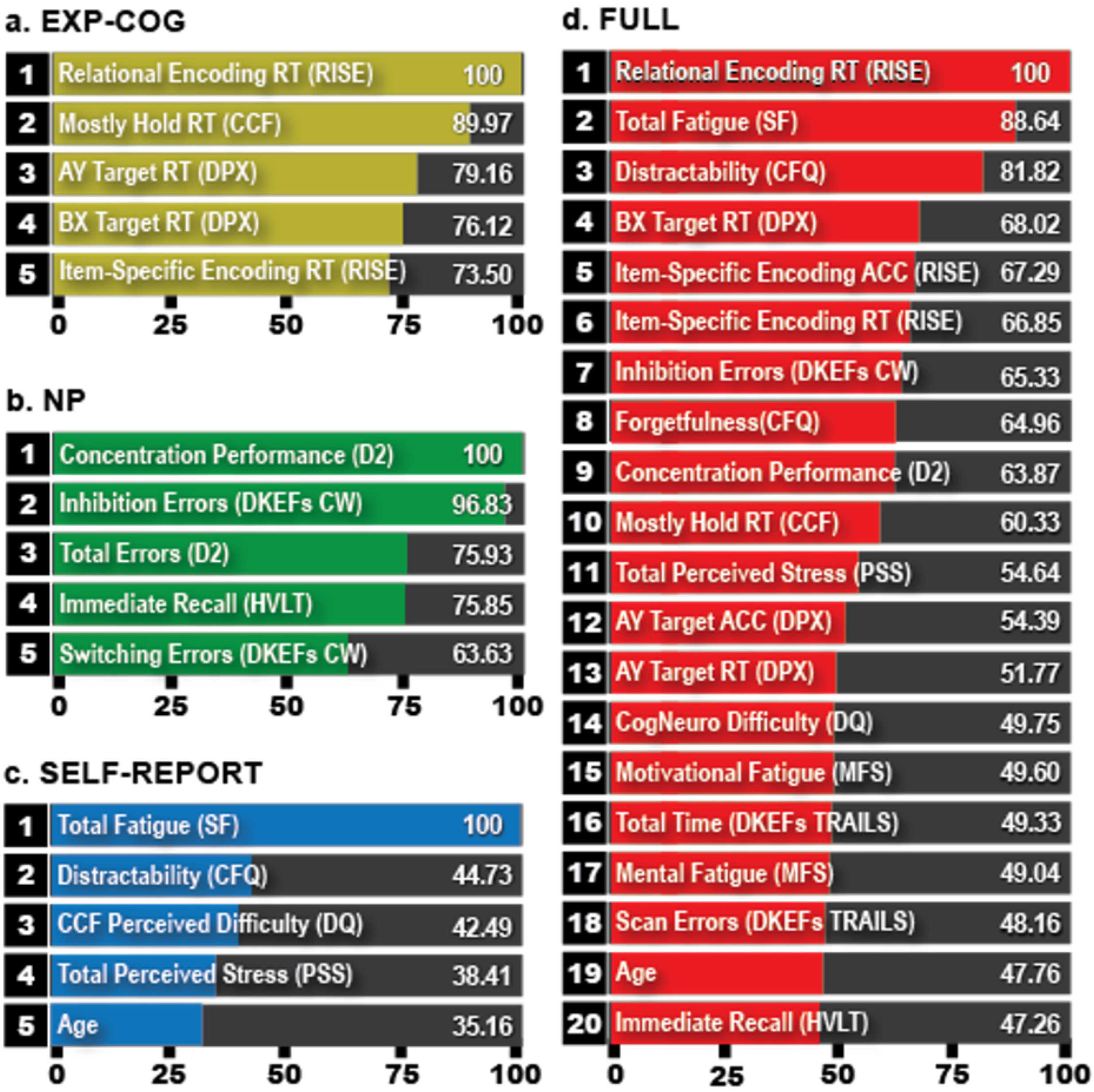


*Note:* Feature importance rankings are displayed for models 1-4. The top 5 features were selected for visualization in the EXP-COG (a), NP (b), and SELF-REPORT (c) models. The top 20 features are depicted for the FULL (d) model.
